# Clinical, biochemical and molecular characterization of newborns with fatty acid β-oxidation disorders: new variants in the *ACADM*, *ACADVL* and *SLC22A5* genes

**DOI:** 10.1101/2025.03.19.25324093

**Authors:** Irene Hidalgo Mayoral, Amanda Herranz Cecilia, Carmen Rodríguez-Jiménez, Ana Carazo Álvarez, Ana Bergua Martínez, José David Andrade Guerrero, Ana Moráis López, Sonia Rodríguez-Nóvoa

## Abstract

**Purpose:** In this study, we aimed to describe clinical, laboratory and molecular features of newborns with clinical suspicion for CTD, MCADD and VLCADD. Fatty acid β-oxidation disorders (FAODs) are a heterogenous group of inherited metabolic defects in which the main common features are fasting or stress-induce hypoketotic hypoglycemia, lactic acidemia or hyperammonemia. The implementation of newborn screening (NBS) programs based on acylcarnitine levels in dried blood spots has allowed them to change the natural course of these diseases, facilitating newborns to be referral to reference units shortly after birth to initiate preventive or therapeutical measures to improve health outcomes. However, handling NBS results is challenging and to date, no correlation between NBS acylcarnitine levels and prognosis has been conclusively established. In this context, genetics plays a key role in identifying individuals with FAODs and in correctly characterizing their disease and their risk of medical complications.

**Methods and results:** This study included 94 newborns who were admitted between 2016 and 2023. Clinical and biochemical data were collected from hospital records. Variant analysis of 21 genes was performed using a custom targeted next generation sequencing (NGS) panel, and variants were classified according to the American College of Medical Genetics and Genomics (ACMG) Standards and Guidelines recommendations. Molecular diagnostic was confirmed in 16/94 (17%) of newborns, and 17 novel variants were detected in in *SLC22A5*, *ACADM* and *ACADVL* genes. We assessed clinical evolution of patients over time and related their clinical status and evolution to their genotype.

**Conclusion:** In this study, we present a retrospective study of 94 newborns with clinical suspicion of CTD, MCADD and VLCADD, and provide clinical, biochemical and genotypic data. We expand the genotypic spectrum of variants in *SLC22A5*, *ACADM* and *ACADVL* and highlight the role of genetics in identifying, characterizing and estimating the risk of medical complications for newborns and its impact on clinical management.

## Introduction

Fatty acid β-oxidation disorders (FAODs) are a heterogeneous group of inherited metabolic defects in fatty acid transport and metabolism that are inherited in an autosomal recessive manner. FAODs occur with an overall estimated rate of incidence of 1:9,300(1). More than 25 genetically distinct disorders have been reported (2), with the most prevalent being medium-chain acyl-CoA dehydrogenase deficiency (MCADD) due to deleterious variants in the *ACADM* gene, systemic primary carnitine deficiency (CTD) due to deleterious variants in the *SLC22A5* gene and very long-chain acyl-CoA dehydrogenase deficiency (VLCADD) due to deleterious variants in the *ACADVL* gene (3).

Fatty acids are the major source of energy under conditions of high metabolic demand, such as prolonged exercise or fasting (4). Therefore, impaired fatty acid oxidation causes depletion of energy levels and can lead to life-threatening metabolic decompensations. Accumulation of non-degraded fatty acids and secondary metabolites in the organism can be monitored(1), being the determination of free carnitine and acylcarnitines values a powerful tool for the diagnosis and monitoring of people with FAODs.

Individuals with FAODs present a highly heterogeneous spectrum in which the main common features are fasting or stress-inducing hypoketotic hypoglycemia, lactic acidemia or hyperammonemia. Most patients remain asymptomatic under appropriate control of intake, although they are not exempt from developing acute, life-threatening metabolic decompensations. The most prevalent symptoms are myocardiopathy, myopathy and liver dysfunction. The age of onset varies according to the disorder, with symptoms such as hypoglycemia being more common in infants whereas others such as rhabdomyolysis occurring more often in older children(5,6).

The implementation newborns screening programs (NBS) for FAODs has allowed to change the natural course of these diseases, limiting their impact in terms of morbidity and mortality. Early detection of FAODs facilitates newborns to be referral to reference units shortly after birth to initiate preventive or therapeutical measures to improve health outcomes(5).

The aim of the present study is to analyze the yield of the NBS program for CTD, MCADD and VLCADD in a single center in terms of diagnosis and monitoring. We present a retrospective study of 94 newborns referred after NBS and describe initial presentation, biochemical and genetic testing and clinical outcomes. We assess the utility of genotyping to resolve the limitations of biochemical diagnosis and to establish clinical prognosis. Furthermore, we provide a set of novel variants in *ACADM*, *ACADVL* and *SLC22A5* genes and assess genotype-phenotype associations.

## Patients and Methods

### 2.1 Patients

Ninety-four newborns with clinical suspicion of β-oxidation disorders were referred to Hospital Universitario La Paz (HULP) in Madrid between 2016 and 2023. Clinical suspicion was based on carnitine or acylcarnitines abnormalities detected through NBS, conducted by tandem mass spectrometry (MS/MS) at the referral laboratory Hospital General Universitario Gregorio Marañón. Blood samples were collected by heel prick in dried blood spots cards at 48h after birth. Primary screening markers for referral were: L-carnitine-(C0) for CTD (reference value or RV = 6.21-43.18 µmol/L), octanoyl-(C8) for MCADD (RV = 0.02-0.13 µmol/L) and cis-5-tetradecenoyl-(C14:1) for VLCADD (RV = 0.04-0.37 µmol/L). Cut-off values are based on the 99.5^th^ percentile levels at 48h of life

All newborns underwent clinical assessment by the clinical monitoring unit of HULP. Medical data of family history, clinical manifestations and treatment were reviewed and retrospectively collected from hospital records.

### 2.2 Genetic studies

Molecular studies were performed at the Metabolic Disease Laboratory of the Genetic Department of HULP. Targeted genes (covering the entire coding regions and exon-flanking sequences) were captured with a custom NGS panel design (SeqCap EZ, Roche) that comprises 742 genes related to metabolic and hepatic disorders. Paired-end sequencing was performed on HiSeq4000 and NovaSeq6000 sequencers (Illumina, San Diego, California, USA). Bioinformatic analysis was conducted by the Bioinformatic Unit using a customized pipeline and NGS data was analyzed using virtual custom panels adapted to the biochemical suspicion, containing a maximum of 21 genes related to fatty acid oxidation and ketonic bodies metabolic disorders (ORPHA:79174(7)): *ACADM, ACADS, ACADVL, ACAT1, CPT1A, CPT2, ETFA, ETFB, ETFDH, FLAD1, HADH, HADHA, HADHB, HMGCL, HMGCS2, MLYCD, OXCT1, SLC16A1, SLC22A5, SLC25A20 and SLC25A32.* Variant prioritization was performed using the following criteria: (i) Minor Allele Frequency, (ii) protein predicted variant impact, (iii) status and ranking of the variants clinical databases, (iv) variant pathogenicity predictors and (v) phylogenetic conservation assessment. Segregation analysis were performed on family members by Sanger sequencing.

Additionally, multiplex-ligation dependent probe amplification (MLPA) was performed to rule out missed copy number variants (CNVs) by NGS. SALSA MLPA Probemix P465 and P076 were used to target *ACADM*, *ACADVL and SLC22A5* genes (MRC-Holland, Amsterdam, The Netherlands). Sequence products were analyzed by capillary electrophoresis (ABI3130XL, Thermo Fisher Scientific). Data was analyzed using Coffalyser.Net software (MRC Holland).

### 2.3 Biochemistry analysis and enzymatic studies

Confirmatory diagnostics for newborn screening and subsequent monitoring studies were conducted at both Centro de Diagnóstico de Enfermedades Moleculares (CEDEM) and Laboratory of Medicine Department of HULP. Acylcarnitines were determined in plasma by liquid chromatography-tandem mass spectrometry (LC–MS/MS) and urine organic acids were determined by LC-MS/MS as trimethylsilyl derivatives. Enzymatic studies were performed in CEDEM. The method used to assess MCAD and VLCAD activity is based on the oxidation of octanoyl-CoA (C8:0-CoA) and palmitoyl-CoA (C16:0-CoA) in the presence of the electron acceptor, ferrocenium hexafluophosphate (FcPF6)(8,9).

## 3. Results

94 newborns showed abnormal acylcarnitine profiles in NBS: ten out of 94 cases (11%) presented low levels of C0 at NBS suggestive of CTD, twenty-five cases (27%) presented increased levels of C8 suggestive of MCADD and fifty-eight cases (62%) presented increased levels of C14:1 suggestive of VLCADD. Summary of referred cases and their final diagnosis is shown in Figure 1.

**Figure 1.**
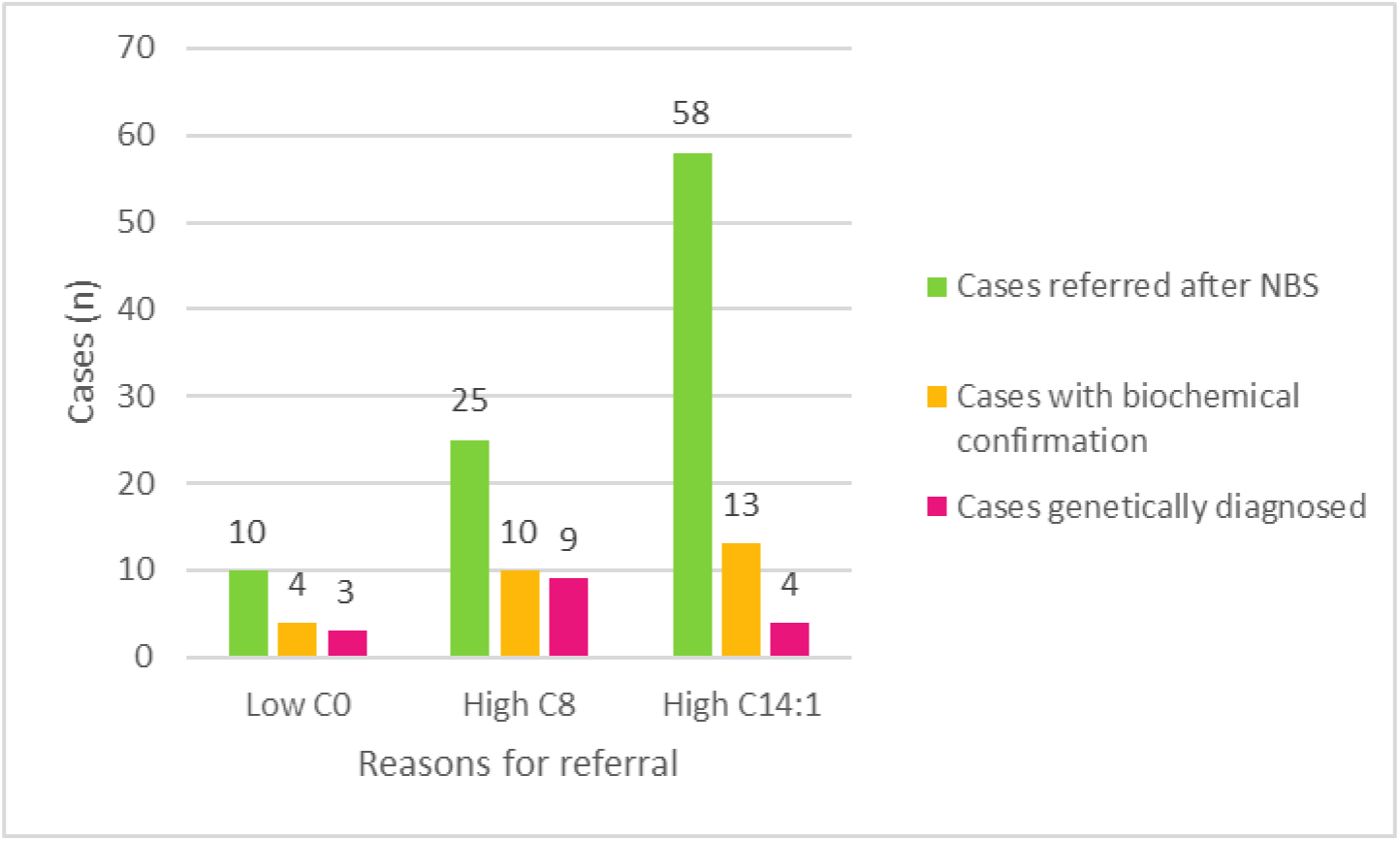
Descriptive distribution of cases among groups. The number of referred cases according to the biochemical suspicion are displayed in yellow. For each group, the number of cases with biochemical confirmation are displayed in blue and the number of cases genetically confirmed are displayed in red.

### 3.1 Suspicion for CTD

Newborns with suspicion for CTD showed median C0 values of 3.575 µmol/L (IR: 6.42-46.21 µmol/L), ranging from 2.12 to 6.07 µmol/L. They were all treated with oral L-carnitine supplementation right after NBS and families were instructed to avoid fasting periods longer than 3-4 hours.

In 4 out of 10 cases (40%), free carnitine levels remained low after the treatment and primary carnitine deficiency was suspected. Diagnosis was confirmed by molecular testing in 3 of the cases, compound heterozygotes for *SLC22A5* deleterious variants, in whom L-carnitine supplementation has been maintained so far (see Table 1 for clinical, biochemical and genetic data). Only one of them (P3) showed metabolic acidosis during episodes of infections. He also showed mild hypotonia and madurative impairment, unlikely related to the disease. The genetically undiagnosed case was heterozygous for a single variant in the *SLC22A5*, and further biochemical testing showed a normalization of C0 levels compatible with a heterozygous carrier status.

**Table 1.** Characteristics of individuals with confirmed molecular diagnosis. ¥ Mother ^(a)^ Compound heterozygosity may not have been confirmed, although it is assumed based on clinical and biochemical data. For cases marked with an asterisk (*), parental testing confirmed that variants were in different alleles. Bold: novel variants ^(b)^ Variant interpretation is based on ACMG criteria or ClinGEN gene-specific guidelines, if available. P: pathogenic, LP: likely pathogenic, VOUS: variant of unknown significance. Variants are referred to reference sequences: NM_003060.4(*SLC22A5*), NM_000016.6(*ACADM*) and NM_000018.4(*ACADVL*) ^(c)^ Acylcarnitine levels from the NBS are displayed numerically when data is available, otherwise they are represented as above (↑) or below (↓) the reference value: C8: VR=0.02-0.13μmol/L; C14:1: VR=0.04-0.38μmol/L; C0: VR=6.42-46.21μmol/L ^(d)^ Reference value for acylcarnitine levels of confirmatory studies vary among laboratories, so results are displayed as times above or below the reference value of each laboratory. ^(e)^ If available, enzyme activity associated with each genotype is reported

| Patient ID | Gene | Genotype<sup>a,b</sup> |  |  |  | NBS<sup>c</sup> | Biochemical confirmatory study<sup>d</sup> | Symptoms at birth | Later symptomatology | Enzymatic activity<sup>e</sup> |
| --- | --- | --- | --- | --- | --- | --- | --- | --- | --- | --- |
|  |  | Variant | Classification | Variant | Classification |  |  |  |  |  |
| P1 | *SLC22A5* | c.760C>T p.(Arg254*) | P | c.1400C>G p.(Ser467Cys) | P | C0=10.77μmol/L | ↓ 0.94 times | No | No | No |
| P2 | *SLC22A5* | c.447C>G p.(Phe149Leu) | VOUS | c.680G>A p.(Arg227His) | LP | C0=5.14μmol/L | ↓ 3.22 times | No | Missed monitoring | No |
| P3* | *SLC22A5* | c.67_69delTTC p.(Phe23del) | P | c.1077T>G p.(Phe359Leu) | VOUS | C0 ↓ | ↓ 2.66 times | No | Metabolic decompensations during infections. Mild hypotonia and maturative impairment | No |
| P4‡ | *SLC22A5* | c.349T>C p.(Trp117Arg) | VOUS | c.529A>G p.(Met177Val) | LP | C0=2.80μmol/L | ↓ 3.89 times (mother) | Not applicable | No | No |
| P5‡ | *SLC22A5* | c.1462C>T p.(Arg488Cys) | LP | c.1462C>T p.(Arg488Cys) | LP | C0=2.94μmol/L | ↓ 6.07 times (mother) | Not applicable | No | No |
| P6‡ | *SLC22A5* | c.149G>A p.(Cys50Tyr) | LP | c.1195C>T p.(Arg399Trp) | P | C0=2.12μmol/L | ↓ 1.29 times (mother) | Not applicable | Cramps and muscle week during pregnancy | No |
| P7‡ | *SLC22A5* | c.34G>A p.(Gly12Ser) | VOUS | c.1403C>T p.(Thr468Ile) | VOUS | C0=2.7μmol/L | ↓ (mother) | Not applicable | No | No |
| P8* | *ACADM* | c.799G>A p.(Gly267Arg) | P | c.985A>G p.(Lys329Glu) | P | C8=4.42μmol/L | ↑ 4.19 times | No | No | No |
| P9 | *ACADM* | c.985A>G p.(Lys329Glu) | P | c.985A>G p.(Lys329Glu) | P | C8=40μmol/L | ↑ 7.31 times | Hypoglycemia<br>Hypotonia | No | No |
| P10* | *ACADM* | c.683C>A p.(Thr228Asn) | P | c.799G>A p.(Gly267Arg) | P | C8=0.5μmol/L | ↑ 1.92 times | No | No | No |
| P11 | *ACADM* | c.985A>G p.(Lys329Glu) | P | c.946-2A>C p.(?) | LP | C8=15.50μmol/L | ↑ 18.022 times | No | Hypoglycemia | No |
| P12 | *ACADM* | c.127G>A p.(Gly267Arg) | P | c.985A>G p.(Lys329Glu) | P | C8=0.38μmol/L | ↑ 3.76 times | No | No | 39% |
| P13* | *ACADM* | c.683C>A p.(Thr228Asn) | P | c.985A>G p.(Lys329Glu) | P | C8= ↑ | ↑ 1.31 times | No | No | 15% |
| P14* | *ACADM* | c.541_552delinsATATC p.(Asp181Ilefs*10) | P | c.683C>A p.(Thr228Asn) | P | C8= ↑ | ↑ 1.53 times | No | No | No |
| P15 | *ACADM* | c.985A>G p.(Lys329Glu) | P | c.985A>G p.(Lys329Glu) | P | C8=7.69μmol/L | ↑ 7.84 times | No | No | No |
| P16 | *ACADM* | c.683C>A p.(Thr228Asn) | P | c.985A>G p.(Lys329Glu) | P | C8=0.69μmol/L | ↑ 1.31 times | No | No | No |
| P17* | *ACADVL* | c.138+2T>C p.(?) | P | c.1366C>T p.(Arg456Cys) | LP | C14:1=1.2μmol/L | ↑ 7.17 times | No | Fever episodes with hyperammonemia | 18% |
| P19 | *ACADVL* | c.853G>C p.(Glu285Gln) | LP | c.1043T>C p.(Leu348Pro) | VOUS | C14:1=5.88μmol/L | ↑ 17.06 times | No | Missed monitoring | No |
| P20* | *ACADVL* | c.358_360del p.(Ala120del) c.956C>T p.(Ser319Leu) | VOUS/VOUS | c.1273G>A p.(Ala425Thr) | LP | C14:1= 5.90 | ↑ 13.37 times | Cardiorespiratory arrest | Rhabdomyolysis, acidosis, gastrostomy carrier. | 4.6% |
| P21* | *ACADVL* | c.305T>C p.(Leu102Pro) | LP | c.538G>A p.(Ala180Thr) | LP | C14:1=7.7μmol/L | ↑ 13.14 times | Bradycardia tend | Rhabdomyolysis and acidosis episodes. Candidate to gastrostomy | 1.5% |

In 6 out of 10 cases (60%), free carnitine levels normalized after L-carnitine treatment and secondary carnitine deficiency was suspected. The mothers of the newborns were tested, showing significant decreased levels of C0 compatible with CTD. Therefore, neonates were considered to have secondary hypocarnitinemia through maternal pass and treatment was ceased. Molecular analysis was performed on the mothers: 4 out of 6 cases (67%) were genetically diagnosed with CTD and started L-carnitine supplementation. Only P7 reported cramps and muscle weakness during previous pregnancy.

Altogether, 14 variants were identified in *SLC22A5* (Figure 2A), including four novel variants: c.189dup p.(Asn64Glnfs*74), c.349T>C p.(Trp117Arg), c.1077T>G p.(Phe359Leu) and c.1403C>T p.(Thr468Ile).

**Figure 2.**
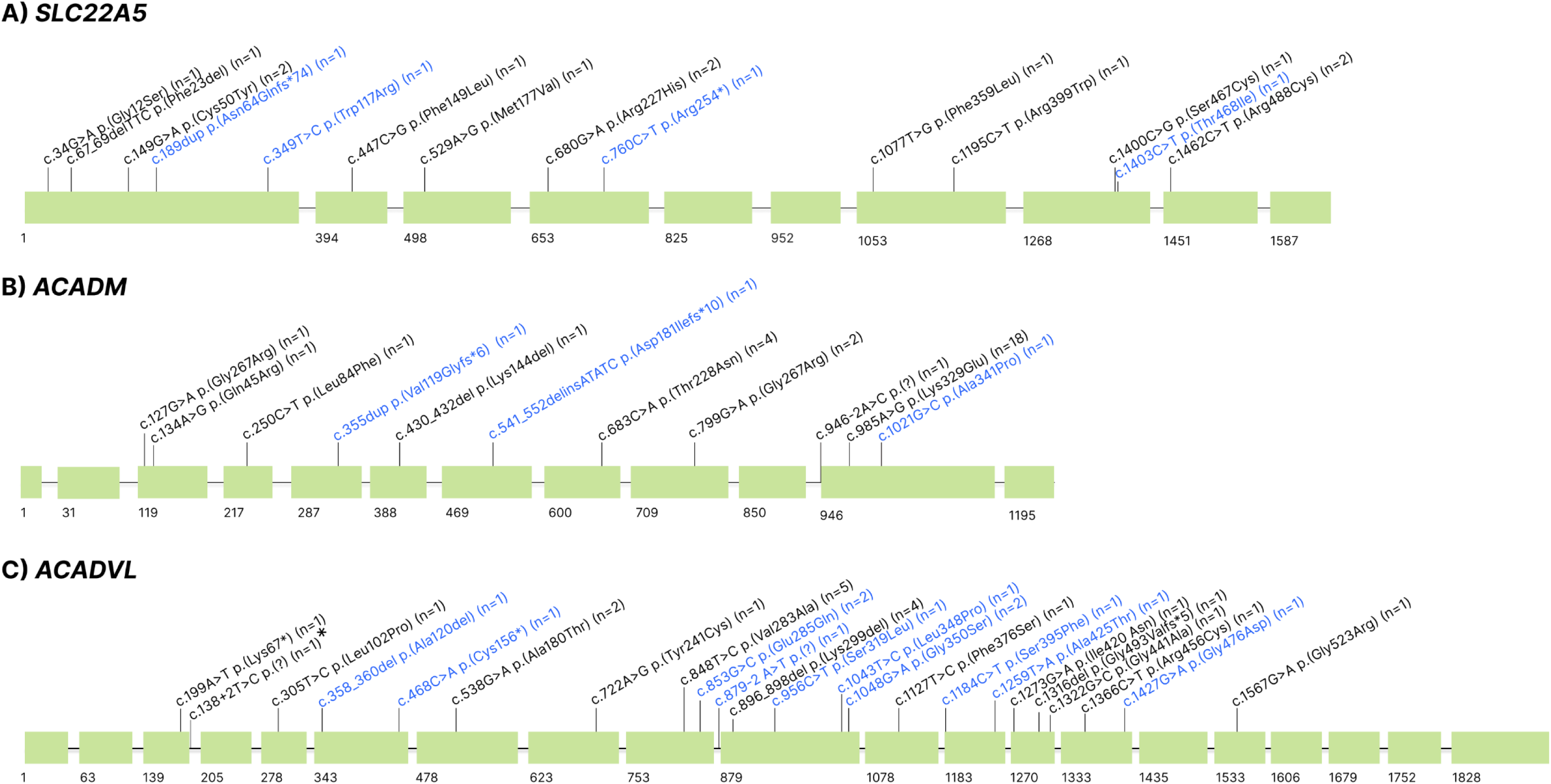
Distribution of variants identified in *SLC22A5*, *ACADM* and *ACADVL*. Representation of genes: boxes represent exons and horizontal lines represent introns. Variants depicted in blue have not been previously reported. Between () are the number of carriers in this cohort. * Previously reported by Martín-Rivada (2022), same patient.

### 3.2 Suspicion for MCADD

Newborns with suspicion for MCADD showed median C8 values of 0.39 µmol/L (IR: 0.02-0.13 µmol/L), ranging from 0.16 to 40 µmol/L. Clinically, only one out of 25 newborns (4%) presented symptoms at birth before NBS results were known (Table 1): P9 presented with symptomatic hypoglycemia (glucose 28 mg/dL, IR: 60-100 mg/dL) that required IV 10% w/v glucose infusion. She presented hypotonia and skin paleness that subsided after glucose normalization.

Breastfeeding or standard infant formula was maintained in all cases after the NBS results, although all families were instructed to avoid fasting periods longer than 3-4 hours and to follow an acting protocol for physiological stress situations before diagnostic confirmation. Biochemical confirmation included in all cases free carnitine and acylcarnitines determination in plasma. In 10 out of 25 cases (40%), C8 levels remained high in confirmatory analysis.

Molecular analysis was performed in all patients with suspicion for MCADD: 9 out of 25 cases (36%) carried homozygous or compound heterozygous variants in the *ACADM* gene and were genetically confirmed, whereas 16 out of 25 cases (64%) carried single variants in *ACADM*. No additional copy number variants were identified in any of the undiagnosed cases. A total of 12 variants were identified in the *ACADM* gene. The complete list of variants is shown in Figure 2B: 8 missense (66%), 2 frameshift (17%), 1 in-frame (8%) and 1 splicing (8.33%). Three of these variants were novel: c.1021G>C p.(Ala341Pro), c.355dup p.(Val119Glyfs*6) and c.541_552delinsATATC p.(Asp181Ilefs*10). The most common variant in our cohort was c.985A>G p.(Lys329Glu), which was identified in 18/25 (72%) newborns (2 homozygotes, 5 compound heterozygotes and 11 heterozygotes).

Confirmed cases of MCADD are presented in Table 1. Nine out of nine cases (100%) showed persistent biochemical abnormalities in confirmatory analysis. All of them are clinical and biochemically monitored, generally once or twice a year. Fluctuating levels of acylcarnitines are observed over time in all cases. However, over the course of this revision, only 1/9 (11%) individuals has developed some symptomatology: patient P11 developed an episode of hypoglycemia (glucose 33 mg/dL, IR: 60-100 mg/dL) during her first years of life. Transitional hypocarnitinemia was seen in 5/9 (56%) children, that required punctual L-carnitine supplementation.

### 3.3 Suspicion for VLCADD

Newborns with suspicion for VLCADD showed median C14:1 values of 0.64 µmol/L (IR: 0.04-0.38 µmol/L), ranging from 0.42 to 7.7 µmol/L (value distribution in Figure 3). Clinically, 8 out of 58 newborns (14%), presented symptoms at birth before NBS results were known. The most common symptom listed was hypoglycemia (n=6), although additional symptoms were reported: hypotonia (n=1), seizures (n=1), hypernatremia with dehydration (n=1) and cardiac involvement (n=2). Two neonates showed cardiac affection: one suffered a cardiorespiratory arrest shortly after birth (P20), whereas the other had a trend towards bradycardia during sleeping (P21) and had a brother history of sudden death shortly after birth.

**Figure 3.**
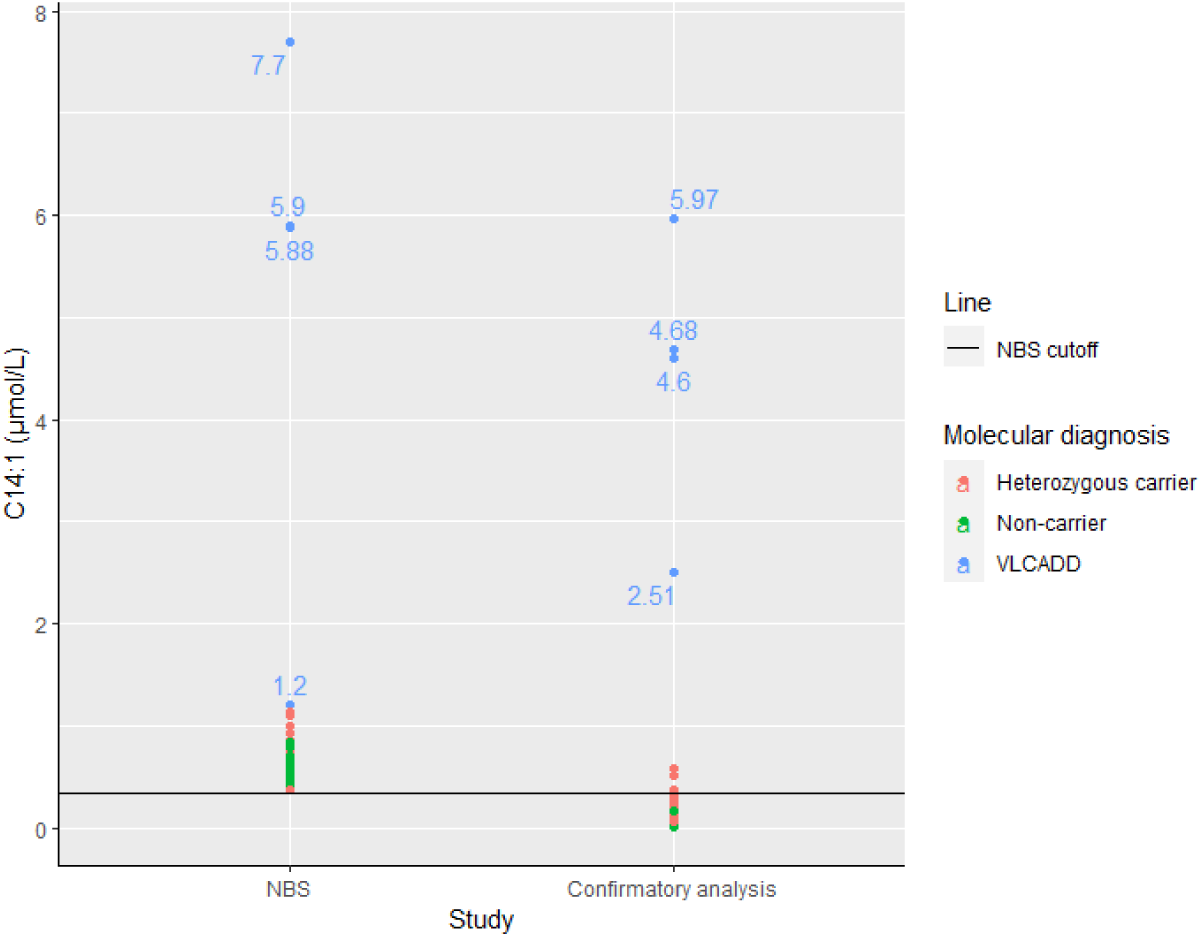
Descriptive distribution of C14:1 values for NBS and confirmatory analysis. NBS cutoff is shown as a horizontal dot line (cutoff <0.35 µmol/L). Confirmatory analysis cutoff is shown as a horizontal plain line. Each value is colored according to the result of the molecular diagnosis (VLCADD confirmed cases are shown in blue, heterozygous carriers in red and non-carriers in green). Values for VLCADD cases are numerically labelled.

All families were instructed to avoid fasting periods longer than 3-4 hours and to follow an acting protocol for physiological stress situations before diagnostic confirmation. Later biochemical confirmation showed that in 13 out of 58 cases (22%), C14:1 levels remained high (Figure 3). In 3 cases breastfeeding was substitute for formulas containing low percentage of long-chain triglycerides (LCT) after NBS results.

Molecular analysis was performed in all patients with suspicion for VLCADD: 4 out of 58 cases (7%) carried compound heterozygous variants in the *ACADVL* gene and were genetically confirmed, whereas 25 out of 58 cases (43%) carried single variants in *ACADVL* and 29 out of 58 cases (50%) carried no variants in *ACADVL*. No additional copy number variants were identified in any of the undiagnosed cases. Altogether, 23 variants were identified in *ACADVL* (Figure 2C): 16 missense (70%), 1 frameshift (4%), 2 nonsense (9%), 2 in-frame (9%), 2 splicing (9%). Among them, 10 variants have not yet been reported in literature: c.358_360del p.(Ala120del), c.468C>A p.(Cys156*), c.853G>C p.(Glu285Gln), c.879-2A>T p.(?), c.956C>T p.(Ser319Leu), c.1043T>C p.(Leu348Pro), c.1048G>A p.(Gly350Ser), c.1184C>T p.(Ser395Phe), c.1259T>A p.(Ile420Asn) and c.1427G>A p.(Gly476Asp).

Confirmed cases of VLCADD are presented in Table 1. Following a positive genetic diagnostic, newborns undergo periodical revisions at the clinical unit that include examination for growth, neurologic and cardiac assessment, and biochemical monitorization. Over the course of this revision, 3/4 (75%) individuals have developed some kind of symptoms: P17 had fever episodes with hyperammonemia and individuals P19 and P20 presented recurrent episodes of rhabdomyolysis and metabolic decompensations episodes with hyperammonemia and hypertransaminasemia, mainly secondary to infections. Both P19 and P20 eventually required gastrostomy-tube placement. Transitional hypocarnitinemia was common and 3/4 (75%) of patients required punctual L-carnitine supplementation. Essential fatty acid deficiency was reported in 3/4 patients who were orally supplemented with arachidonic and docosahexaenoic acid.

## 4. Discussion

Herein, we report a retrospective study of the results and impact of the NBS program for CTD, MCADD and VLCADD in a single center in Madrid in terms of diagnosis and management. All variants identified in this study have been registered to ClinVar under the accession numbers SCV005423831--SCV005423860. Molecular diagnostic was confirmed in 16 out of 94 individuals (17%): 9 cases of MCADD, 4 cases of VLCADD and 3 cases of CTD. Diagnostic rates varied among clinical suspicion groups, ranging from 3/10 (30%) in newborns with low C0 levels to 9/25 (36%) in newborns with increased C8 levels and to 4/58 (7%) in newborns with increased initial C14:1 levels (Figure 1). The remaining cases include 43 heterozygous carriers (46%) and 31 cases with no variants identified (33%), that no longer needed to be under supervision. Additionally, 4 mothers were diagnosed of unaware CTD, increasing the diagnosis yield to 20 out of 94 cases in this cohort (21%).

### Individuals with suspicion for CTD

Interpreting C0 levels of NBS requires caution, since isolated low C0 levels may be observed both in primary CTD (due to a defect in the OCTN2 transmembrane transporter of L-carnitine) and secondary CTD. Secondary deficiencies due to maternal transfer have been described in other clinical entities such as 3-methylcrotonyl-CoA carboxylase deficiency(10) or glutaric aciduria type I(11) and it has been widely described in *CTD*(12,13). An approach for discriminating primary and secondary CTD consists in treating newborns with oral carnitine and measuring their response. Those cases in which C0 levels remain low are suggestive of a primary defect whereas in cases in which C0 levels normalize, free NBS carnitine levels reflect those of the mother shortly after birth(14).

In this cohort, 3 newborns were diagnosed with CTD due to compound heterozygous missense variants in the *SLC22A5* gene that codifies the widely expressed OCTN2 transporter. Only P1 carried a truncating variant, c.760C>T p.(Arg254*), and interestingly, P1 showed the highest free carnitine levels at NBS. Treated newborns are usually asymptomatic since manifestations can be prevented by maintaining normal plasma carnitine levels(15). All our cases were asymptomatic except case P3 that carried the genotype c.[149G>A];[1195C>T], who suffered metabolic decompensations during infections. Additionally, 4 maternal CTD cases were unmasked, all of them asymptomatic over years except P7 who reported cramps and muscle weakness during her last pregnancy. CTD in untreated adults may lead to fatigability or absence of symptoms, although symptoms may manifest or exacerbate during pregnancy. NBS constitutes thus a powerful tool for unmasking asymptomatic women who may benefit of close monitoring of plasma carnitine concentrations and increasing their carnitine supplementation to cover body requirements during pregnancy(16).

In this study, we identified 4 novel variants in *SLC22A5*: c.189dup p.(Asn64Glnfs*74), c.349T>C p.(Trp117Arg), c.760C>T p.(Arg254*) and c.1403C>T p.(Thr468Ile). Two of them, c.189dup p.(Asn64Glnfs*74) and c.760C>T p.(Arg254*) are truncating variants that codify a OCTN2 transporter that lacks more than half of its sequence or that may lead to its degradation by nonsense-mediated decay.

### Individuals with suspicion for MCADD

MCADD is the most prevalent disease in our cohort. In Spain, the estimated prevalence is 1:17.480(17). It shows the highest diagnostic rates after NBS, with a positive predictive value (PPV) of 36%. Following a positive genetic diagnostic, newborns underwent periodical examination and biochemical monitorization. Families were instructed to avoid fasting periods longer than 4 hours during the day and longer than 3 to 12 hours during the night, depending on the child’s age. No drastic diet changes were made, although hypocaloric diets containing less than 30% of total energy from fat are prescribed and products with medium-chain triglycerides (MCT) should be avoided(18).

False positives may be due to low weight or physiological stress at birth and medication. Additionally, heterozygous carriers of variants in *ACADM* have slightly increased levels of C8 that normalize after weeks(19). Confirmatory testing was consistent: 60% of cases normalize C8 levels and all of them turned out to be heterozygous carriers after genetic analysis. Later confirmatory analysis of C8 or using secondary biomarkers such as C8/C10 ratios can help to reduce false positives. On the other hand, 40% of cases remain high in confirmatory testing, and among those only one case was finally not diagnosed of MCADD by molecular testing. The non-diagnosed case presented C8 levels slightly above the reference limit and further evaluation was consistent with a carrier-MCADD status, ruling out the possibility of missing the diagnosis.

MCADD is mainly caused by deleterious missense variants in the *ACADM* gene that disrupt MCAD oligomerization and leads to its loss-of-function through protein misfolding(20). In this study we identified three novel variants in the *ACADM* gene: c.355dup p.(Val119Glyfs*6), c.541_552delinsATATC p.(Asp181Ilefs*10) and c.1021G>C p.(Ala341Pro). Two of these variants, c.355dup p.(Val119Glyfs*6) and c.541_552delinsATATC p.(Asp181Ilefs*10) create a truncated protein that lacks 70% and 55% of its sequence, respectively. Thus, critical regions for MCAD catalytic activity or proper oligomerization may be lost. The third novel variant, c.1021G>C p.(Ala341Pro), is an unknown significance variant that may disrupt MCAD oligomerization and lead to MCAD misfolding(20), although further analysis are required to establish its role in MCAD loss-of-function.

The most frequent variant is c.985A>G p.(Lys329Glu), that accounts for 90% of defective alleles reported in literature. In this cohort, variant c.985A>G p.(Lys329Glu) was present in 7 out of 9 diagnosed newborns: cases P9 and P15 carried a homozygous genotype whereas cases P8, P11, P12, P13 and P16 carried a compound heterozygous genotype. Genotype has been reported to correlate with residual enzyme activity: the homozygous genotype is associated with very low residual enzyme activity while compound heterozygous individuals have higher residual enzyme activity, varying from 11 and 30%(8). We did not test MCAD activity for homozygous cases P9 and P15, although they presented significantly higher C8 levels at NBS (P9: 40 μmol/L and P15: 7.69 μmol/L) than compound heterozygous, consistent with a less degree of enzymatic activity. The only exception is case P11 (C8=15.50 μmol/L), compound heterozygous for the c.985A>G p.(Lys329Glu) and the c.946-2A>C p.(?) variants. Variant c.946-2A>C has been described to cause skipping of exon nine(21), where c.985A>G is located, and therefore it may mimic a homozygous state for c.985G>A variant that would explain C8 levels at NBS for this patient. In our study, decompensations were only observed in individuals P9 and P11, presenting hypoglycemia episodes at birth and overtime that were resolved without further complications. Our findings support correlation between higher C8 newborn screen values, homozygosity for the c.985A>G and the risk for clinical complications, previously described(22,23).

On the other hand, heterozygous genotypes for the c.985A>G p.(Lys329Glu) are associated with better prognosis(24,25) and none of the children presented any metabolic decompensation nor symptoms until now. MCAD activity was tested for cases P12 and P13, showing a residual enzyme activity of 39% and 15% respectively, consistent with the expected milder phenotype.

### Individuals with suspicion for VLCADD

Elevated levels of C14:1 were the main cause of referral in this cohort, although after molecular analysis, the final diagnosis of VLCADD was achieved only in 6.8% of patients. The high false-positive rate observed is related to the fact that long-chain acylcarnitines levels tend to be high at 48h after birth, leading to a positive screening, and then tend to normalize within a few days. False positive results may also be associated with treatments, catabolic situations like prolonged fasting between maternal feeding, milder phenotypes or severe body weight loss(21,22). Additionally, it has been reported that heterozygous carriers of variants in *ACADVL* have slightly increased levels of C14:1 that are related to the low specificity of the initial NBS assay(26–28). Several strategies may increase NBS specificity such as C14:1 ratio(29,30), urine dicarboxylic acids(31) or new biomarkers like 3-4-dihydroxytetradecanoylcarnitine(32), although they are not yet implemented in routine healthcare diagnostic flows.

Later biochemical confirmatory analysis seems to be a powerful tool to stratify more accurately the risk for VLCADD of newborns. In 78% of cases, confirmatory testing showed normal levels of C14:1, suggestive of non-carrier or heterozygous-carrier condition. Families were instructed to avoid fasting periods longer than 4 hours during the day, but breastfeeding was not changed. In 22% cases C14:1 levels remained high, and after genetic testing 4 were molecularly diagnosed with VLCADD whereas the remaining 9 turned out to be heterozygous carriers. VLCADD cases presented C14:1 values above 7 times the upper limit of the reference value: low-fat formula containing low long-chain triglycerides (LCT) were introduced and families were instructed to avoid fasting periods longer than 3-4 hours. Nutrition therapy for individuals with VLCADD may vary depending on the severity of the phenotype: symptomatic and severe forms are typically treated by restricting long-chain fatty acids and increasing the percentage of MCT of the diet, whereas in moderate and mild forms breastfeeding is allowed, but complemented with low LCT formulas(33). Additionally, newborns were referred to cardiological units for clinical evaluation. On the contrary, heterozygous carriers presented C14:1 values less than 4 times the upper limit of the reference value and although families were instructed to avoid fasting, no dietary fat composition change was strictly prescribed at the beginning.

C14:1 levels vary among heterozygous carriers and VLCADD cases, suggesting a correlation with the disease. Although we didńt observe the overlap of biochemical analytes in confirmatory testing in our cohort, genetic testing still needs to be performed to accurately discriminate both situations and to safely label unaffected non-risk newborns that no longer need to be under supervision. Genotyping is a powerful tool to resolve pitfalls in VLCADD diagnosis and to guide clinical management of newborns after a positive NBS result(34).

After genetic testing, more than 20 unique variants in *ACADVL* gene have been identified in this cohort, including eleven novel variants. Up to 75% of them were substitutions, being the most frequent the c.848T>C p.(Val283Ala) variant. It accounts for nearly 30% of defective alleles reported in literature(35) and decreases the enzyime activity to 20-25%(36). VLCAD is a homodimer of 67 KDa subunits anchored to the inner mitochondrial membrane. Substitutions may induce changes in substrate or cofactor location by disturbing the hydrophobic environment, preventing crucial contacts or inducing conformational changes that modify the substrate binding cavity or the homodimer aggregation(37).

Despite VLCADD is the less common disorder in our cohort, individuals seem to be at a higher risk of developing medical complications with a wide spectrum of severity. Individual P17 carried the c.[138+2T>C];[1366C>T] genotype, associated to a residual enzyme activity of 18%. She reported no cardiac nor muscular complications so far, although she has presented hyperammonemia during fever and infections episodes, normalizing after glucose intravenous infusion. On the other hand, individuals P20 and P21 carried genotypes related to worse clinical courses, suggesting their association with a higher degree of enzymatic deficiency that was confirmed by enzymatic studies in lymphocytes: P20 carried the c.[358_360del;956C>T];[1273G>A] genotype (residual activity: 4.6%) whereas P21 carried the c.[305T>C];[538G>A] genotype (residual activity:1.5%). Both genotypes seem to lead to a higher risk of metabolic decompensations over time, requiring gastrostomy placement to manage poor oral feeding and prevent hypoglycemia. Use of gastrostomy feedings is a widely used strategy for individuals with severe forms of VLCADD who are not able to maintain metabolic control by oral feeding, especially throughout the night(33). Additionally, they have developed multiple rhabdomyolysis episodes over time (CK levels >1000 UI/L), which is a frequently reported long-term complication of VLCADD(34). No cardiac complications have been observed over the years, although both genotypes seem to be associated with a higher risk for cardiological symptomatology: P20 suffered a cardiorespiratory arrest right after birth and P21 showed bradycardia tend during the newborn period. P21 had an older brother that died of sudden death shortly after birth, carrying the same genotype. Thus, genotype c.[305T>C];[538G>A] may be associated with cardiac involvement. Cardiomyopathies are common in patients with VLCADD, presenting a variable spectrum that includes dilated cardiomyopathy, hypertrophic cardiomyopathy and cardiac arrhythmias. Although they can be asymptomatic, cardiological monitorization is indicated given the high risk of sudden infant death syndrome(38).

## 5. Conclusion

CTD, MCADD and VLCADD increase the risk of developing acute, life-threatening metabolic decompensations. NBS has allowed early detection of newborns at risk, facilitating their referral to reference units shortly after birth to initiate preventive or therapeutical measures. However, its low specificity with the actual cutoffs and lead to a high number of false positives that turn out to be heterozygous or non-carrier individuals who dońt require especial management. Given the difference among groups in terms of clinical management, it is essential to accurately being able to determine the individuaĺs risk of having a FAOD. Nevertheless, no correlation between *NBS* acylcarnitine levels and prognosis have been conclusively established to date and there is overlapping of acylcarnitine levels with the actual cutoffs. In this respect, genetics plays a key role both in identifying individuals with FAODs and in correctly characterizing their disease and their risk of medical complications. This work contributes to expanding the genotypic spectrum of variants in *SLC22A5*, *ACADM* and *ACADVL* and to delineating clinical and biochemical consequences of certain genotypes.

## Statements and declarations

## Author contributions

Irene Hidalgo Mayoral: conceptualization, data curation, formal analysis, writing-original draft. Amanda Herranz Cecilia: data curation, formal analysis, writing-review and editing. Carmen Rodríguez-Jiménez: methodology, writing-review and editing. Ana Carazo Álvarez: methodology, writing-review and editing. Ana Moráis López: formal analysis, writing-review and editing. Ana Bergua Martínez: formal analysis, writing-review and editing. José David Andrade Guerrero: formal analysis, writing-review and editing. Sonia Rodríguez-Nóvoa: conceptualization, writing-review and editing.

## Data availability

The datasets generated and/or analysed during the current study are available on request from the corresponding author. The data are not publicly available due to privacy or ethical restrictions. All variants are publicly available on ClinVar database.

## Ethics approval

The Ethics Committee of ‘La Paz’ University Hospital in Madrid, in accordance with the Declaration of Helsinki, approved the present study (PI-6086).

## Consent to participate

Informed consent for the use of patient’s data, materials and/or test results for research purposes was included in the written informed consent, which was obtained before sample collection

